# Hepatitis C transmission and progression among people who have injected drugs in Singapore: Modelling treatment for eradication

**DOI:** 10.1101/2025.10.24.25338708

**Authors:** Minah Park, Yichen He, Shihui Jin, Nigel W H Lim, Jue Tao Lim, Si Jack Chong, Borame L Dickens

## Abstract

**Background:** Hepatitis C virus (HCV) disproportionally affects people who inject drugs (PWID), posing a public health challenge. The high incarceration rate among PWID provides an opportunity for screening and treating individuals with chronic HCV infection in correctional facilities.

**Methods:** We developed a deterministic age-structured model to characterise the HCV transmission among PWID both inside and outside prison settings. We proposed diverse treatment strategies, targeting varying proportions of current and former PWID. Utilising demographical and epidemiological data on PWID in Singapore, we assessed the impacts of these strategies on preventing HCV infections and HCV-related complications.

**Results:** The overall HCV prevalence among PWID is estimated at 31.3% (95% CrI: 30.7% – 32.1%) without intervention. Treating all detained ex-PWID in the early stages of chronic HCV infection could prevent 25,878 infections (95% CrI: 20,485 – 31,575) over 50 years, comparable to treating 15% of current PWID in the community. However, treating 15% of former PWID in the community would have a greater impact in reducing advanced HCV cases and HCV-related deaths. A combined approach covering 12% of current PWID, all detained ex-PWID, and 13% of former PWID in the community would nearly eliminate HCV infections among PWID, reducing infections to 0.3% (95% CrI: 0.25% – 0.45%) of ever-PWID 30 years after treatment initiation.

**Interpretation:** Eliminating HCV infections among PWID is achievable but costly, particularly when targeting all PWID regardless of incarceration. Treating detained PWID offers a more feasible and cost-effective alternative. Policymakers should strike a balance between budget constraints and treatment effectiveness to optimise public health outcomes.

## 1. INTRODUCTION

Hepatitis C virus (HCV) infection is a leading cause of liver-related illness and deaths. Despite continued efforts, viral hepatitis remains a significant public health concern, with the number of attributable deaths steadily increasing over the past decades. Globally, nearly 400,000 people are estimated to die each year from HCV-related complications, including cirrhosis and hepatocellular carcinoma [1, 2]. Of an estimated 71 million people living with chronic HCV infection worldwide in 2015, 14 million were from the Western Pacific Region, where there were an estimated 110,000 people newly infected with HCV [2].

A bloodborne virus, HCV, is commonly transmitted through sharing contaminated needles, placing people who inject drugs (PWID) at higher risk of the infection. Current evidence suggests that PWID account for approximately 23% of new HCV infections and 33% of HCV-related deaths worldwide [1]. While there is no effective vaccine against HCV to date, research has shown that antiviral medicines such as direct-acting antivirals (DAA) are highly effective in clearing the virus, with a cure rate of at least 90% for patients with chronic HCV infection in 12 weeks [3]. In the absence of vaccines, early diagnosis and DAA treatment may be the most effective means to prevent and reduce the burden of HCV-related illness.

Yet, utilisation of DAA therapies remains low due to limited access to HCV screening coupled with the high cost of DAAs. The World Health Organisation (WHO) estimated that only 19% of those infected with HCV knew about their infection status, and 15% of those diagnosed received treatment by 2017 [1]. Such a low uptake of HCV testing and treatment among at-risk populations impedes the public health effort to eliminate HCV infection, and it is important to ensure that they are routinely screened for HCV to receive timely treatment. Implementing community-wide interventions to promote HCV testing among PWID remains challenging, however, as they are often marginalised and stigmatised in society.

Considering high incarceration rates among PWID and high HCV prevalence in correctional settings [4], offering HCV testing and treatment to the detained PWID has great potential to reduce the prevalence not only in prisons but also in the community. Numerous studies found such a prison-based, test-and-treat strategy to be cost-effective in the UK and the US, where routine periodic testing for all PWID is recommended by the US Centres for Disease Control and Prevention [4–7].

In the Western Pacific Region, Australia has recently expanded the national HCV screening and treatment scheme, focusing on people who have injected drugs, including those in correctional facilities [8]. Yet, compared to other major infectious diseases, HCV is relatively underexplored in the region, and more robust efforts are required at the national level to join the global effort to eliminate HCV infection (i.e., 90% and 65% reduction in incidence and mortality, respectively) by 2030 [9].

Therefore, we aimed to examine the effectiveness of offering DAA treatment to priority populations, including (i) PWID, (ii) ex-PWID in the community and (iii) those in correctional facilities. Specifically, we developed a mathematical model of HCV transmission, progression, and treatment to measure the impact of various treatment strategies on the burden of HCV among PWID of different age groups in Singapore.

## 2. METHODS

Using a deterministic age-structured model, we simulated various treatment strategies in which a different group of populations is treated at varying proportions. Key outcomes of the study included the prevalence of HCV infections, the number of HCV-related complications such as hepatocellular carcinoma (HCC), the number of individuals to be treated under these different treatment scenarios, and the number of deaths among patients with advanced HCV.

### 2.1 Data source

The demographic and epidemiological information of all detained PWID admitted to Changi Prison between 2007 and 2017 was provided by the Singapore Prison Service (SPS). Age-specific prevalence of HCV was derived from the results of the universal screening (i.e., non-reactive, borderline-reactive, or reactive) conducted among 5,086 new and returning detained PWID between December 2014 and February 2016.

### 2.2 Model of incarceration and infection

The dynamic model consisted of three states to reflect incarceration and drug use status (D: never or formerly detained PWID, J: detained ex-PWID, X: formerly detained ex-PWID) and eight states for HCV infection status (Figure 1). Each compartment was further split into nine age groups of 5 years (15–19, 20–24, 25–29, 30–34, 35–39, 40–44, 45–49, 50–54, and 55+), with individuals entering the model at age 15–19 as uninfected, current PWID. In addition, ageing was modelled to capture transitions of current and ex-PWID into older age groups over time, with mortality applied at age-specific rates from the Singapore Department of Statistics [10].

**Figure 1.**
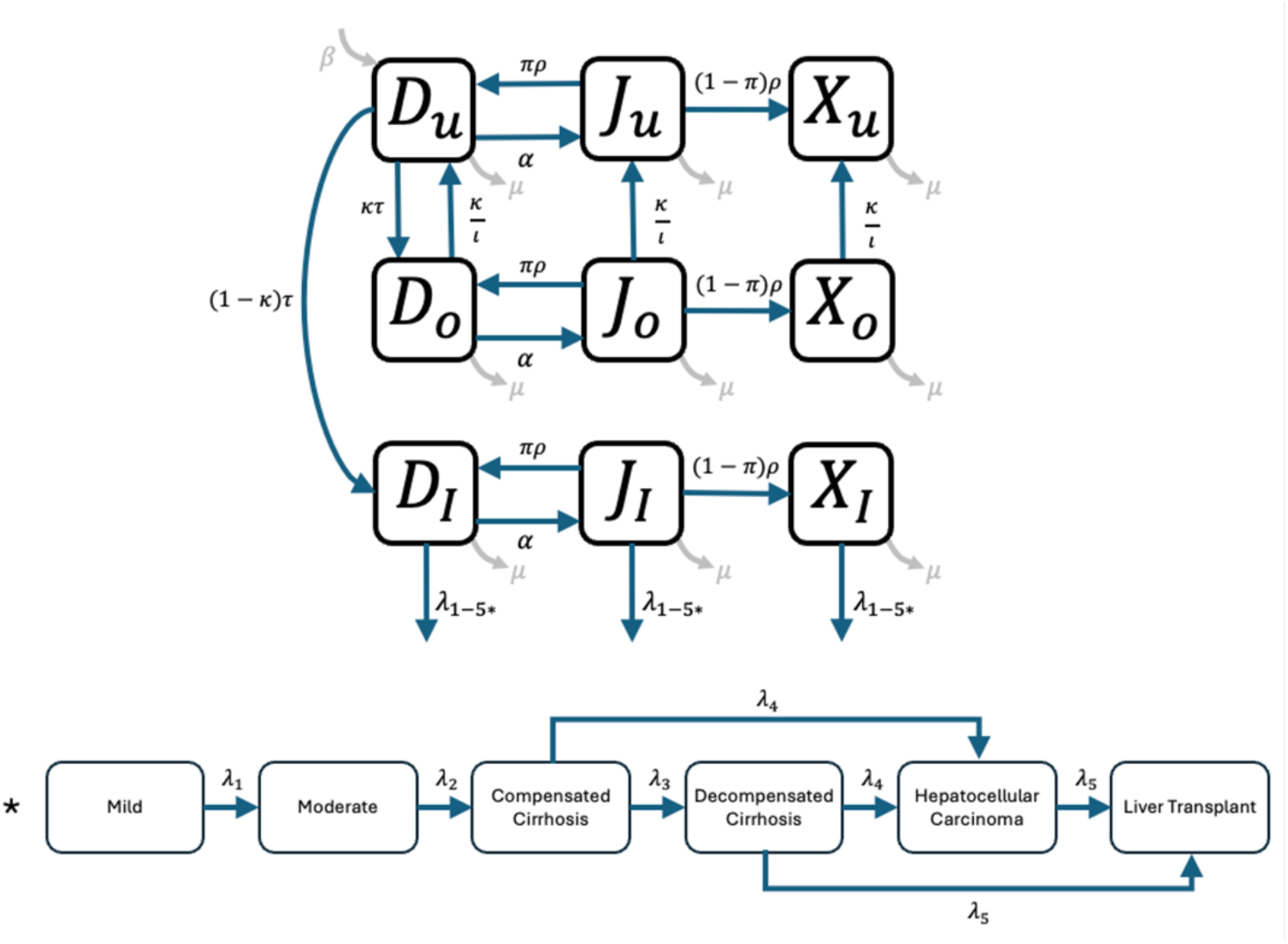
Simplified model schematic for incarceration dynamics and HCV transmission. All state variables are time dependent, with the temporal parameter omitted for simplicity. Flow between incarceration states is shown in the diagram above, while disease progression following infection is presented in the diagram below. The compartments *D* refer to never or formerly detained PWID, *J* the detained population of PWID and *X* the formerly detained ex-PWID. The subscripts *u* refer to those uninfected, *o* acute infections that spontaneously clear and *I* the infection pathway from mild HCV infection to terminal stages where a liver transplant is eventually required. DAA treatment was assumed to be available to individuals with mild and moderate HCV infection and those with compensated cirrhosis. Please refer to Table 1 for details on parameters.

For age group *i* (∈ {1,2, ⋯,9}), let 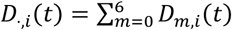 denote the number of infected drug users, *D*_*u,i*_(*t*) the number of uninfected drug users, and *N*_⋅,*i*_(*t*) = *D*_*u,i*_(*t*) + *D*_⋅,*i*_(*t*) the number of all drug users.

**Table 1.** Parameters and states in the model, with values where estimates exist.

| Quantity | Interpretation | Value | Source |
| --- | --- | --- | --- |
| $D_{u,i}(t)$ | Current PWID, uninfected with HCV<br>($i = 1, 2, 3, \dots, 9$) | Time- and age-varying | |
| $D_{j,i}(t)$ | Current PWID, infected with HCV ($i = 1, 2, 3, \dots, 9$; $j = 1, 2, 3, \dots, 6$) | Time- and age-varying | |
| $J_{u,i}(t)$ | Detained ex-PWID; uninfected | Time- and age-varying | |
| $J_{j,i}(t)$ | Detained ex-PWID; infected | Time- and age-varying | |
| $X_{u,i}(t)$ | Formerly detained ex-PWID; uninfected | Time- and age-varying | |
| $X_{j,i}(t)$ | Formerly detained ex-PWID; infected | Time- and age-varying | |
| $n(t)$ | Total population size | Time- and age-varying | |
| $\beta$ | Initiation rate, per year | 1,861 | SPS |
| $a_i$ | Arrest rate per current drug user per year | Age-varying | SPS |
| $\pi_i$ | Recidivism probability, per detained former user per incarceration | Age-varying | SPS |
| $\rho_i$ | Release rate per detained former user per year | Age-varying | SPS |
| $m_{i,j}$ | Average number of needle sharing events between age groups $i$ and $j$ | Age-varying | [11] |
| $p_i$ | Scaling factor for contact-based force of infection | Age-varying | Fitted |
| $\tau$ | Transmission probability, per needle sharing event | 0.0057 | [12] |
| $\kappa$ | Proportion of infections leading to spontaneous clearance | 0.26 | [13] |
| $l$ | Duration in which spontaneous clearers are infectious | 180 days | [13] |
| $\lambda_1$ | Transition probabilities from mild to moderate HCV per year | 0.025 | [6] |
| $\lambda_2$ | Transition probabilities from moderate to cirrhosis per year | 0.037 | [6] |
| $\lambda_3$ | Transition probabilities from cirrhosis to decompensated cirrhosis per year | 0.039 | [6] |
| $\lambda_4$ | Transition probabilities from cirrhosis or decompensated cirrhosis to HCC per year | 0.014 | [6] |
| $\lambda_5$ | Transition probabilities from decompensated cirrhosis or HCC to liver transplant per year | 0.03 | [6] |
| $\theta_1$ | Sustained viral response (SVR) for those with mild and moderate HCV | 0.97 | [14] |
| $\theta_2$ | SVR for those with compensated cirrhosis | 0.94 | [14] |
| $\mu_i$ | Mortality rate, per person per year | Age-varying | [10] |
| $\mu^4$ | Mortality rate per year, decompensated cirrhosis to death | 0.13 | [6] |
| $\mu^5$ | Mortality rate per year, HCC to death | 0.43 | [6] |
| $\mu^6$ | Mortality rate per year, LT to death | 0.21 | [6] |

The number of new infections for age group *i* at time *t* would be

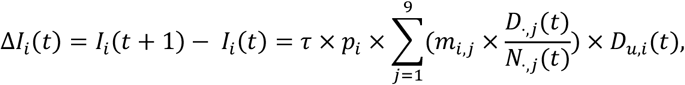

where *τ* is the transmission probability per needle sharing event, *p*_*i*_ is age-specific scaling factors for contact-based force of infection estimated using the observed prevalence data within a Bayesian framework, and *M* = (*m*_*i,j*_) is a 9 × 9 contact matrix representing the average number of needle sharing events between age groups *i* and *j*, sourced from a study of PWID in Vietnam due to the absence of local empirical data [11]. Disease progression among infected individuals followed pre-defined transition rates^6^ and was assumed independent of release and recidivism. Mortality rates were elevated for individuals with decompensated cirrhosis, HCC, and liver transplant states to reflect liver disease severity. All state variables, epidemiological and prison parameters are shown in Table 1.

We further made the following assumptions: i) PWID would be detained at age-dependent rates; ii) Upon release from prison, some would return to taking drugs while others would never again take drugs; iii) Sojourn times were independent and exponentially distributed, with inter-arrest intervals following the same distribution as that from initiation of drug use to first arrest; iv) Only current PWID would be at risk of acquiring or transmitting HCV through sharing infected needles; and v) Spontaneous clearance would occur in 26% of the acutely infected individuals within the first 6 months of infection,^10^ with the remainder progressing to six stages of chronic infection defined by fibrosis severity. Further details of the model structure and fitting procedures are provided in the Supplementary Information.

### 2.3 HCV Treatment

We assumed that current and ex-PWID diagnosed with mild infection to compensated cirrhosis would be treated for 12 weeks using oral DAA with a cure rate of 94% and 97% [14]. Following the Hepatitis C Guidance 2019, individuals with decompensated cirrhosis and further complications were assumed not to receive DAA treatment due to little improvement in clinical outcomes [15]. Of treated individuals, those who have successfully achieved sustained viral response (SVR) were assumed to not progress to the next stage, but to remain susceptible to reinfection. The untreated or those not responsive to the treatment (non-SVR) were, however, assumed to move to the next stage of HCV infection. Treatment was assumed to take immediate effect following 12 weeks of treatment.

As outlined in Table 2, each intervention scenario was designed to treat a different group of the population by varying respective proportions. Treatment was assumed to occur once a month. The model was run for baseline and five intervention scenarios over a time window of 50 years.

**Table 2.** Summary of treatment scenarios used in the model.

| Strategy | Treatments given to: |
| --- | --- |
| 0 | None (Baseline) |
| 1 | Proportion of current PWID (1% – 15%) |
| 2 | All currently detained ex-PWID* (2a – 2c) |
| 3 | Proportion of formerly detained ex-PWID (1% – 15%) |
| 4 | Combined (12% current PWID + all currently detained ex-PWID and 10% –13% formerly detained ex-PWID) |

\*: individuals with compensated cirrhosis (2a), moderate or compensated cirrhosis (2b) or mild to compensated cirrhosis (2c)

The combined strategy, in which a fraction of all three groups were treated, was considered to estimate the minimum number of treatments required to reach the goal of HCV elimination among PWID within 30 years of intervention. A one-way sensitivity analysis was performed, where SVR rates were reduced by 20% for active drug users, to explore the impact on the effectiveness of the various treatment strategies above.

## 3. RESULTS

### 3.1 HCV prevalence, recidivism and release rates among PWID

Detained PWID aged 40–44 years were the most likely to resume infection drug use within 10 years upon release, while those aged 55 years and above had the lowest recidivism probability. Elder detained PWID, particularly those aged 45–54 years, tended to spend more time in prison compared to other age groups, with a median duration of 2.39 years for individuals aged 35 years or above, and 1.77 years for those under 35 (Figure S1). Approximately 60% of the drug users detained during the study period were repeat offenders, and a detained PWID was expected to experience 2.78 lifetime incarcerations. The typical total prison time was 5.91 years, averaging 2.12 years per incarceration.

The overall prevalence of HCV infection among the screened detained PWID was estimated at 31.3% (95% credible interval [CrI]: 30.7% – 32.1%). The baseline prevalence was inferred to increase with age, rising from 18.1% (95% CrI: 16.5% – 19.4%) among young offenders aged 15–19 years, peaking at 44.0% (95% CrI: 42.4% – 45.6%) for those aged 45–49 years, and subsequently declining slightly to 40.0% (95% CrI: 38.4% – 41.6%) among individuals aged 55 years and above (Figure S2). These estimates are generally consistent with the observed data, although minor discrepancies in the 40–44 and 55+ age groups may reflect unaccounted-for cohort effects.

### 3.2 Effectiveness of diverse treatment strategies in averting HCV infections

Treating either current PWID or the detained ex-PWID could substantially reduce the prevalence of HCV infection in the next 50 years (Figure 3). Targeting the current PWID and treating 15% of those with mild to compensated cirrhosis would potentially lower the total number of infections to under 10,000 by year 50, with counts among PWID or those in prison dropping to 136 (95% CrI: 61 – 349) or lower. Focusing on the detained individuals would yield a similar effect when those with up to compensated cirrhosis were involved in treatment, with slightly fewer cases to be averted among former PWID in the community (by 1,044, 95% CrI: 321 – 2,113) and fewer reductions among current PWID (by 1,015, 95% CrI: 179 – 4,851) by year 50. In contrast, treating formerly detained ex-PWID would not impact the prevalence among current PWID or former PWID in prison, but it would significantly reduce HCV infections among the former PWID in the community to nearly 5,000 cases within 20 years. It is also worth noting that the strategy offering treatment to all the three groups (12% of PWID, all detained ex-PWID, and 13% of the formerly detained ex-PWID diagnosed with mild to compensated cirrhosis) could reduce the number of active HCV infections to 590 (95% CrI: 521 – 866) within 30 years of intervention (Figure 3).

**Figure 3.**
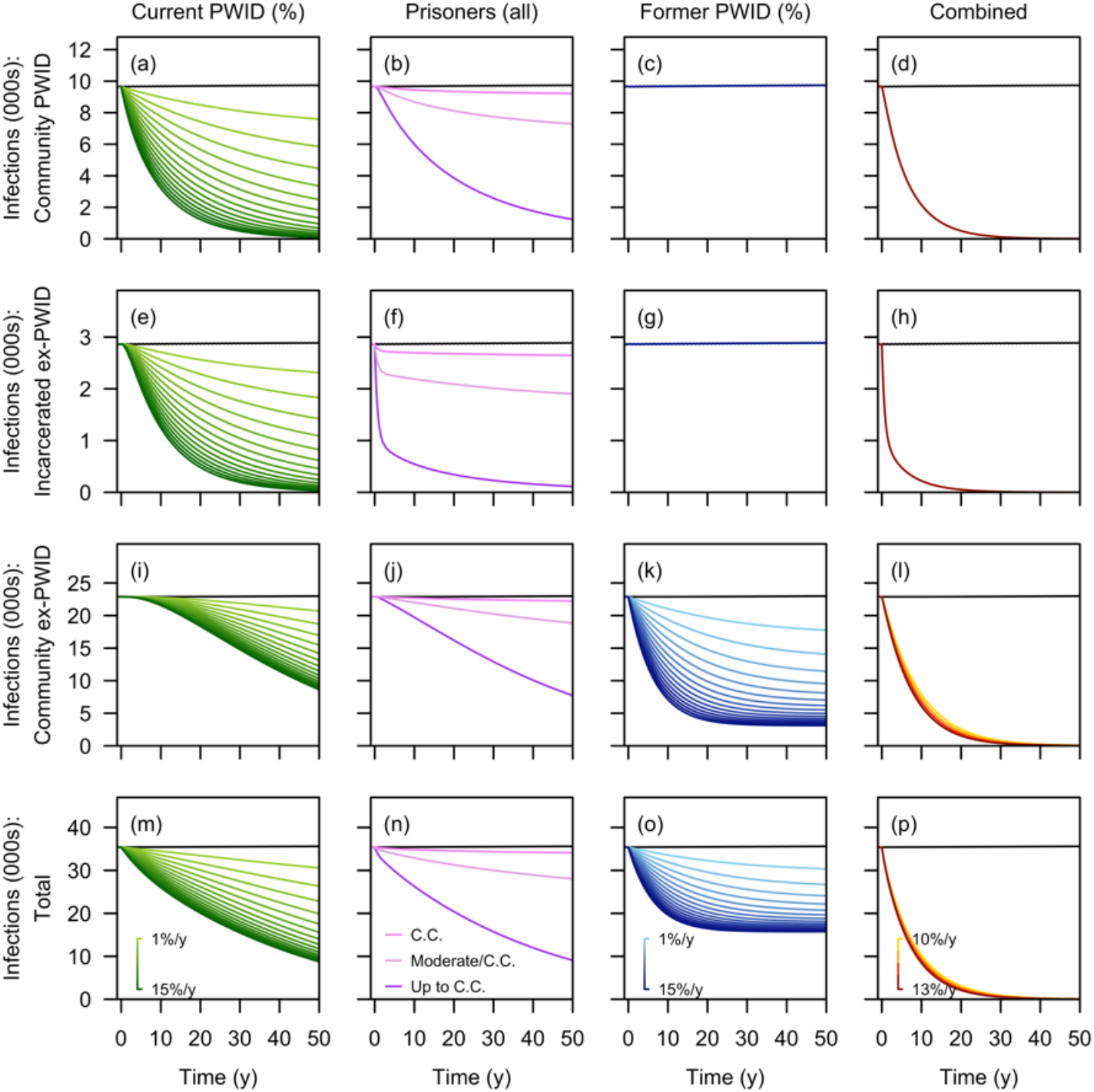
Projected numbers of HCV infections under various treatment scenarios over 50 years. These scenarios include treating a fraction of current PWID (Column 1), inmates at specific infection stages (Column 2), a fraction of former PWID (Column 3), and a combined approach involving 12% of current PWID, all inmates, and a fraction of former PWID (Column 4). Outcomes of each treatment scenario covering different population sizes are indicated by colour-coded lines, with darker lines representing broader coverage. The black horizontal line in each panel shows the baseline scenario with no treatment. The four rows are for infection counts among current PWID (Row 1), detained ex-PWID (Row 2), released ex-PWID in the community (Row 3), and the overall population (Row 4).

In addition, treating these priority groups with early stages of chronic HCV infection also helps prevent complications such as decompensated cirrhosis and hepatocellular carcinoma. Interventions prioritising formerly detained ex-PWID would result in the greatest reduction in HCV-related complications. The strategy, however, would require more financial and healthcare resources, as it would require triple the number of treatments required compared to the strategies targeting current PWID or detained ex-PWID. The combined strategy remains the most effective and efficient in averting HCV-related complications. When the targeted treatment group includes 12% of current PWID, all ex-PWID in prison, and 13% of former PWID in the community, this approach would substantially lower the annual number of new cases with decompensated cirrhosis and HCC to 14 (95% CrI: 14 – 16) and 3 (95% CrI: 3 – 4), respectively, within the first 20 years of intervention. Its impact on averting deaths among ever PWID in advanced HCV stages was also the most prominent among all proposed treatment plans, with up to 275 deaths prevented per year. Although this scheme would also require the most resources in the initial 20 years, the number of individuals needing treatment would become minimal with the gradual elimination of the disease (Figure 4).

**Figure 4.**
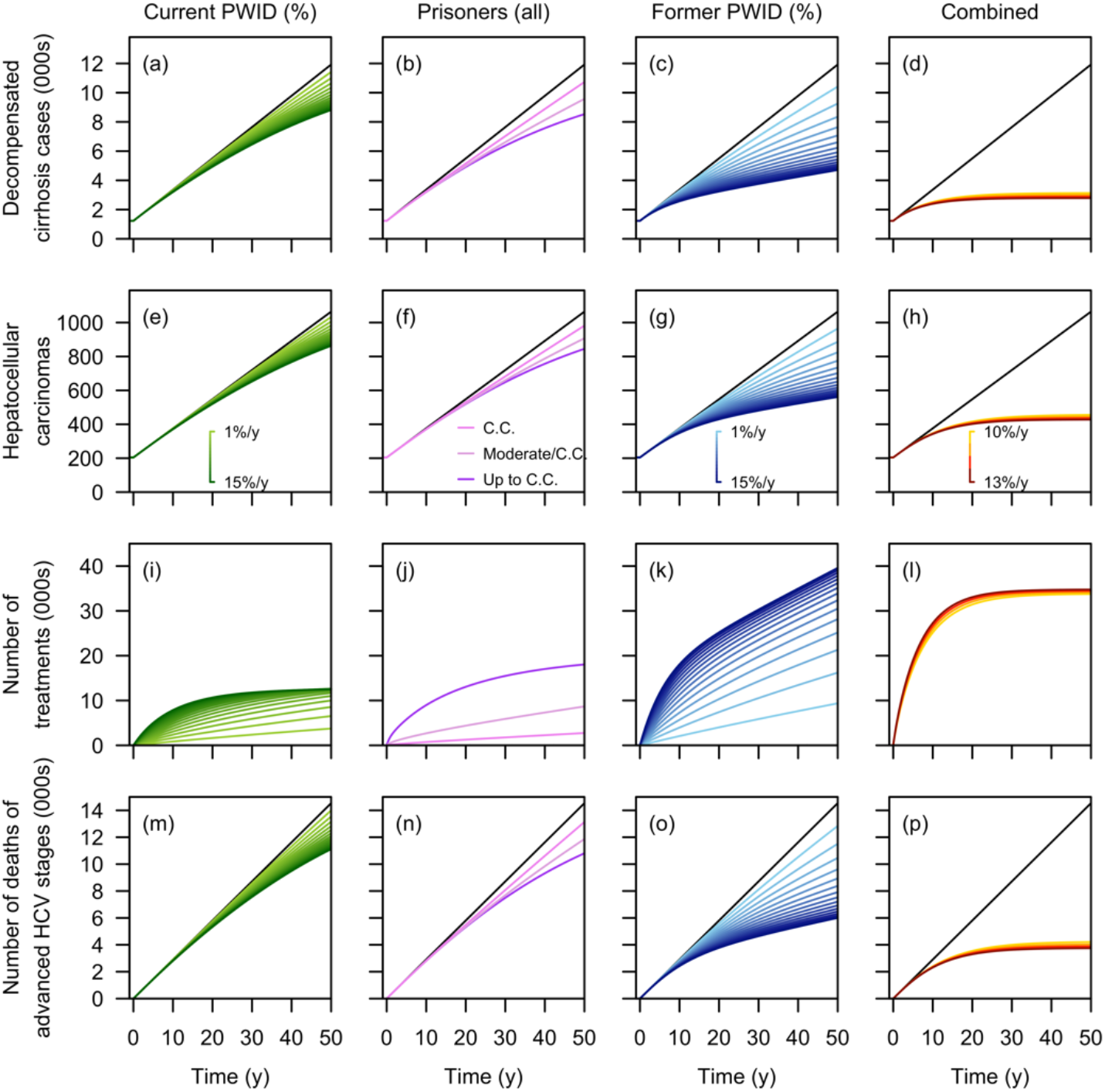
Projected cumulative number of current and former PWID with advanced HCV stages, the corresponding treatments, and deaths over 50 years. The modelled scenarios include treating a fraction of current PWID (Column 1), inmates at specific infection stages (Column 2), a fraction of former PWID (Column 3), and a combined approach involving 12% of current PWID, all inmates, and a fraction of former PWID (Column 4). Outcomes of each treatment scenario covering different population sizes are indicated by colour-coded lines, with darker lines representing broader coverage. The black horizontal line in each panel shows the baseline scenario with no treatment. The three rows are for the number of individuals with decompensated cirrhosis (Row 1), hepatocellular carcinomas (Row 2), the number of treatments required (Row 3), and total deaths among individuals in advanced HCV stages (Row 4).

### 3.3 Projected HCV prevalence among current and ex-PWID by year 30

Targeting all ex-PWID in prisons, or 15% of current or former PWID in the community could effectively reduce the prevalence to 2.40% (95% CrI: 1.12% – 4.85%), 1.53% (95% CrI: 1.16% – 2.27%), and 2.54% (95% CrI: 2.10% – 3.13%) within their respective groups over 30 years of intervention. The prevalence in the total population, however, would decrease to 9.38% (95% CrI: 8.31% – 11.52%), 9.23% (95% CrI: 8.89% – 9.84%), and 9.34% (95% CrI: 7.21% – 12.38%), under these three strategies, respectively. The combined approach—involving all PWID in prison, 12% of current PWID, and 13% of former PWID in the community who were infected with HCV and had mild to compensated cirrhosis— offers the most promising path to eliminating HCV infection, lowering the proportion of the HCV infections among ever PWID to below 0.5% (Table 3).

**Table 3.**
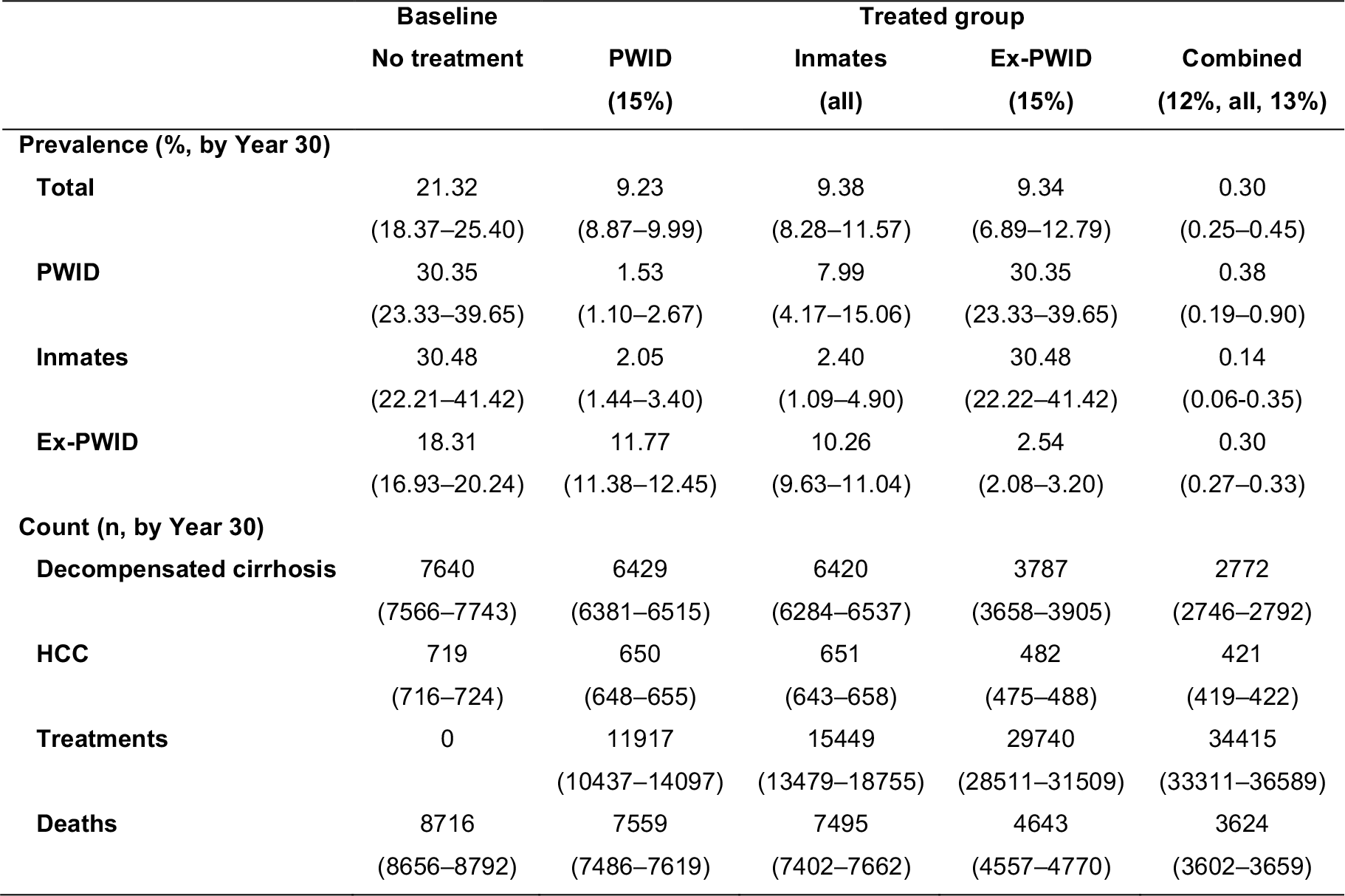
Estimated prevalence and cumulative number of HCV-related complications, treatments required, and deaths among individuals in advanced HCV stages under selected treatment scenarios by year 30.

The sensitivity analysis demonstrated the robustness of our results to variations in the cure rates of the hypothetical DAAs. Reducing the SVR rate by 20% for current PWID as compared to ex-PWID had minimal impact on the overall prevalence and the number of treatments required, resulting in a minor 2.46% (95% CrI: 0.64%-4.97%) change over the 30-year time horizon (Table S2).

## 4. DISCUSSION

In this study, we established a dynamic, age-stratified model of incarceration, HCV transmission, progression, and treatment among current and former PWID. We proposed and evaluated a range of treatment strategies, demonstrating the high effectiveness of treating all detained PWID diagnosed with early stages of chronic HCV infection in reducing the overall HCV burden, with up to 15,000 infections averted over 50 years. The modelled results indicate that the implementation of HCV screening in prison settings could lower HCV prevalence among both current and detained PWID, with subsequent reductions in infection rates and longer-term health complications.

Our results also indicate that fewer courses of treatment would be required to achieve the same effectiveness when focusing on current PWID instead. Limited awareness of HCV, however, combined with social stigma and fear of discrimination [16], often discourages individuals from seeking screening and treatment, making it challenging to meet the treatment goal of 15% of PWID, as simulated in our analysis. By comparison, a prison-based intervention is more feasible, as over half of the ever-PWID have a history of incarceration, allowing easier access to the target treatment group [17]. Furthermore, such an intervention may also complement existing community-based outreach programmes aimed at reducing HCV transmission among active drug users transitioning in and out of prisons [18].

While treating formerly detained ex-PWID in the community has minimal impact on HCV prevalence in PWID or those in prison, and hence on curbing transmission, it emerges as the most effective strategy for reducing the incidence of decompensated cirrhosis and HCC. This approach significantly reduces the societal burden by decreasing morbidity and mortality attributable to HCV, primarily affecting older ex-PWID, particularly those aged 55 years and above (Table S1) who face a higher risk of progressing to more advanced stages of HCV infection and developing severe complications [1]. In our simulations, with a treatment coverage rate of 15%, the projected number of treatments required per year would decrease exponentially, starting from around 2600 in the first year and stabilising at approximately 450 annually from year 20 onwards. Provided the high costs of DAAs and constrained healthcare budgets [19], this strategy may not be practical, especially if prevalence reduction is the primary goal.

However, provided sufficient budget allocation for HCV treatment and a commitment to HCV elimination among PWID, the combined strategy is highly recommended. This approach, which encompasses all the ever-PWID subgroups, could significantly reduce the infection rates among active PWID or those in prison to nearly zero, while circumventing the suboptimal downstream impacts of focusing solely on these two groups in terms of sequelae and mortality. Furthermore, this strategy’s potential to eliminate HCV within 30 years suggests a promising future with little disease burden and reduced investment in treatment thereafter, making it a more cost-effective solution in the long run.

Our analysis has limitations. These include the assumption of static rates for injection initiation and HCV infection rates over the 50-year simulation window, which may vary over time due to shifts in drug use patterns and needle-sharing behaviours [20]. We relied on estimates from the literature [11] and approximations of empirical incarceration statistics provided by SPS to inform key parameters. Yet, there remains uncertainty as to the true values of these quantities, which we addressed through Monte Carlo methods in addition to sensitivity analyses. Similarly, while we used progression rates of HCV infection from a UK-based study [6], which are plausible for biological processes, country-specific factors, such as available healthcare resources and liver transplant rates [21], may limit the direct applicability of these rates to our context.

Nonetheless, while HCV prevalence among detained PWID remains high at 30% without interventions, both this group and the prevalence among active drug users could be substantially lowered to below 3% in 30 years with sufficient investment in treatment for all the PWID in prison with mild to compromised cirrhosis. The potential for HCV elimination remains feasible if continuity of care can be ensured while individuals transit in and out of prisons, as demonstrated by the treatment effects of the combined strategy targeting a wider population of both current and former PWID, regardless of their incarceration status.

To sum up, our assessment of various treatment strategies for averting HCV infections demonstrates the possibility of eliminating HCV among ever-PWID in an Asia Pacific prison setting. The prioritisation of treatment for detained PWID could be considered due to the higher disease prevalence in this population, easier accessibility, and comparable treatment effectiveness in reducing disease prevalence. Disease elimination would be more efficiently achieved with a comprehensive approach targeting all individuals with a history of drug injection, though this would necessitate greater resource investment in the initial decade of implementation. Policymakers should therefore carefully weigh the trade-off between the required investment and the overall effectiveness of treatment strategies to maximise the benefit to human well-being within a limited budget.

## Data Availability

All data produced in the present study are available upon reasonable request to the authors

## List of abbreviations

DAA: direct-acting antivirals
HCC: hepatocellular carcinoma
HCV: Hepatitis C virus
PWID: people who inject drugs
SPS: Singapore Prison Service
SVR: sustained viral response
WHO: World Health Organisation
UK: United Kingdom
US: United States

## Declarations

### Ethics approval and consent to participate

Ethical review and informed consent were waived for this study as it involved analysis of anonymised secondary data provided by Singapore Prison Service. Ethics exemption was sought through SSHSPH DERC RNR process.

### Author contributions

M.P. and B.L.D. conceived and designed the study. S.J.C. compiled and curated the data. M.P. and Y.H. implemented the statistical analysis and created the figures and tables. M.P. and Y.H. wrote the original draft of the manuscript. S.J., N.W.H.L., S.J.C., J.T.L., and B.L.D. reviewed and edited the manuscript.

### Data and code availability

The analytical codes for this study can be accessed at https://github.com/Mister-He/HepC-Transimission-Modelling. The raw data on the detained PWID admitted to Changi Prison are available upon reasonable request.

## Acknowledgements

Not applicable.

