## Supplementary Information for "Hepatitis C transmission and progression among people who have injected drugs in Singapore: Modelling treatment for eradication"

### Model Specification

In this study, we built a deterministic compartmental model, with the population stratified into nine age groups. At each time $t$, the state variables tracked the populations of drug users ($D$), prisoners ($J$), ex-prisoners ($X$) as well as cumulative number of deaths and number of treats. The infection state for each subpopulation was further categorized into acute (spontaneously resolving) or chronic (including mild, moderate, compensated cirrhosis, decompensated cirrhosis, hepatocellular carcinoma and liver transplantation). The detailed transition between different states are as follows:

### Infection

The number of new infections in age group $i$ at time $t$ was determined by disease prevalence in each subpopulation and age-specific mixing across groups:

$$\Delta I_i(t) = I_i(t+1) - I_i(t) = \tau \times p_i \times \sum_{j=1}^{9} (M_{i,j} \times \frac{D_{\cdot,j}(t)}{N_{\cdot,j}(t)}) \times D_{u,i}(t),$$

with the corresponding force of infection being

$$\gamma_i(t) = \tau \times p_i \times \sum_{j=1}^{9} (M_{i,j} \times \frac{D_{\cdot,j}(t)}{N_{\cdot,j}(t)}).$$

Here, $p_i$ is age-specific scaling factors for contact-based force of infection, $\tau$ is the transmission probability per needle sharing event, and $M$ is a $9 \times 9$ contact matrix representing the average number of sharing events between age groups $i$ and $j$.

### Never or formerly detained PWID ($D$)

New initiatives entered the youngest susceptible age group (15–19, $D_{u,1}$) at rate $\beta$. Together with their older counterparts, they were exposed to infection at an age-specific force of infection $\gamma_i(t)$. A fraction ($\kappa$) was assumed to clear the infection spontaneously within duration $\iota$, while the rest developed chronic HCV infection, progressing from mild to more advanced stages ($D_{1:6,i}$) with fixed transition rates ($\lambda_{1:5}$). Meanwhile, at each time step, individuals in age group $i$ were subject to arrest at rates $\alpha_i$, and following release with probability $\rho_i$, a proportion $\pi_i$ of ex-prisoners rejoined the PWID population. Mortality was modelled with age-specific rates $\mu_i$, and higher rates applied for advanced liver disease stages ($\mu^4, \mu^5, \mu^6$):

$$
\begin{aligned}
\frac{d}{dt}D_{u,i} &= -\gamma_i(t)D_{u,i} + \pi_i\rho_i J_{u,i} - \alpha_i D_{u,i} + \frac{\kappa}{\iota}D_{0,i} - \mu_i D_{u,i} + 1_{i=1}\beta,\\
\frac{d}{dt}D_{0,i} &= \kappa\gamma_i(t)D_{u,i} + \pi_i\rho_i J_{0,i} - \alpha_i D_{0,i} - \frac{\kappa}{\iota}D_{0,i} - \mu_i D_{0,i},\\
\frac{d}{dt}D_{1,i} &= (1-\kappa)\gamma_i(t)D_{u,i} + \pi_i\rho_i J_{1,i} - \alpha_i D_{1,i} - \lambda_1 D_{1,i} - \mu_i D_{1,i},\\
\frac{d}{dt}D_{2,i} &= \lambda_1 D_{1,i} + \pi_i\rho_i J_{2,i} - \alpha_i D_{2,i} - \lambda_2 D_{2,i} - \mu_i D_{2,i},\\
\frac{d}{dt}D_{3,i} &= \lambda_2 D_{2,i} + \pi_i\rho_i J_{3,i} - \alpha_i D_{3,i} - (\lambda_3+\lambda_4)D_{3,i} - \mu_i D_{3,i},\\
\frac{d}{dt}D_{4,i} &= \lambda_3 D_{3,i} + \pi_i\rho_i J_{4,i} - \alpha_i D_{4,i} - (\lambda_4+\lambda_5)D_{4,i} - \mu^4 D_{4,i},\\
\frac{d}{dt}D_{5,i} &= \lambda_4 D_{4,i} + \pi_i\rho_i J_{5,i} - \alpha_i D_{5,i} - \lambda_5 D_{5,i} - \mu^5 D_{5,i} + \lambda_4 D_{3,i},\\
\frac{d}{dt}D_{6,i} &= \lambda_5 D_{5,i}v - \mu^6 D_{6,i} + \lambda_5 D_{4,i},
\end{aligned}
$$

Detained ex-PWID ($\boldsymbol{J}$)

The transitions between different disease states for detained ex-PWID were similar to those for never or formerly detained PWID, but were further influenced by incarceration and release flows:

$$
\begin{aligned}
\frac{d}{dt}J_{u,i} &= \alpha_i D_{u,i} - \pi_i\rho_i J_{u,i} - (1-\pi_i)\rho_i J_{u,i} + \frac{\kappa}{\iota}J_{0,i} - \mu_i J_{u,i},\\
\frac{d}{dt}J_{0,i} &= \alpha_i D_{0,i} - \pi_i\rho_i J_{0,i} - (1-\pi_i)\rho_i J_{0,i} - \frac{\kappa}{\iota}J_{0,i} - \mu_i J_{0,i},\\
\frac{d}{dt}J_{1,i} &= \alpha_i D_{1,i} - \pi_i\rho_i J_{1,i} - (1-\pi_i)\rho_i J_{1,i} - \lambda_1 J_{1,i} - \mu_i J_{1,i},\\
\frac{d}{dt}J_{2,i} &= \alpha_i D_{2,i} - \pi_i\rho_i J_{2,i} - (1-\pi_i)\rho_i J_{2,i} + \lambda_1 J_{1,i} - \lambda_2 J_{2,i} - \mu_i J_{2,i},\\
\frac{d}{dt}J_{3,i} &= \alpha_i D_{3,i} - \pi_i\rho_i J_{3,i} - (1-\pi_i)\rho_i J_{3,i} + \lambda_2 J_{2,i} - (\lambda_3+\lambda_4)J_{3,i} - \mu_i J_{3,i},\\
\frac{d}{dt}J_{4,i} &= \alpha_i D_{4,i} - \pi_i\rho_i J_{4,i} - (1-\pi_i)\rho_i J_{4,i} + \lambda_3 J_{3,i} - (\lambda_4+\lambda_5)J_{4,i} - \mu^4 J_{4,i},\\
\frac{d}{dt}J_{5,i} &= \alpha_i D_{5,i} - \pi_i\rho_i J_{5,i} - (1-\pi_i)\rho_i J_{5,i} + \lambda_4 J_{4,i} - \lambda_5 J_{5,i} - \mu^5 J_{5,i} + \lambda_4 J_{3,i},\\
\frac{d}{dt}J_{6,i} &= \lambda_5 J_{5,i} - \mu^6 J_{6,i} + \lambda_5 J_{4,i}.
\end{aligned}
$$

Formerly-detained ex-PWID ($\boldsymbol{X}$)

Formerly-detained ex-PWID also followed similar infection and disease progression processes, with additional release and recidivism dynamics explicitly incorporated:

$$\begin{aligned}
\frac{d}{dt}X_{u,i} &= (1-\pi_i)\rho_i J_{u,i} + \frac{\kappa}{\iota}X_{0,i} - \mu_i X_{u,i},\\
\frac{d}{dt}X_{0,i} &= (1-\pi_i)\rho_i J_{0,i} - \frac{\kappa}{\iota}X_{0,i} - \mu_i X_{0,i},\\
\frac{d}{dt}X_{1,i} &= (1-\pi_i)\rho_i J_{1,i} - \lambda_1 X_{1,i} - \mu_i X_{1,i},\\
\frac{d}{dt}X_{2,i} &= (1-\pi_i)\rho_i J_{2,i} + \lambda_1 X_{1,i} - \lambda_2 X_{2,i} - \mu_i X_{2,i},\\
\frac{d}{dt}X_{3,i} &= (1-\pi_i)\rho_i J_{3,i} + \lambda_2 X_{2,i} - (\lambda_3+\lambda_4) X_{3,i} - \mu_i X_{3,i},\\
\frac{d}{dt}X_{4,i} &= (1-\pi_i)\rho_i J_{4,i} + \lambda_3 X_{3,i} - (\lambda_4+\lambda_5) X_{4,i} - \mu^4 X_{4,i},\\
\frac{d}{dt}X_{5,i} &= (1-\pi_i)\rho_i J_{5,i} + \lambda_4 X_{4,i} - \lambda_5 X_{5,i} - \mu^5 X_{5,i} + \lambda_4 X_{3,i},\\
\frac{d}{dt}X_{6,i} &= \lambda_5 X_{5,i} - \mu^6 X_{6,i} + \lambda_5 X_{4,i}.
\end{aligned}$$

#### Mortality

Let $M_{Y,s,i}(t)$ denote the cumulative number of deaths up to time $t$, for incarceration state $Y$ ($D$ for never detained PWID, $J$ for detained ex-PWID, and $X$ for former-detained ex-PWID), disease stage $s \in \{u, 0, \cdots, 6\}$, and age group $i \in \{1, \cdots, 9\}$. The increase in the death counts was:

$$\frac{d}{dt}M_{Y,s,i} = \mu_i Y_{s,i}.$$

#### Treatment

Treatment was assumed to occur once per month, following the completion of daily transitions and applied to individuals in the early disease stages ($s = 1,2,3$) across all compartments. Coverage was determined by the state-specific proportions ($p_{D_s}$, $p_{J_{s,\cdot}}$ and $p_{X_s}$) specified for each strategy (Table 2). Let $Y_{s,i}^-$ and $Y_{s,i}^+$ ($Y \in \{D, J, X\}$) denote population before and after treatment, $\theta_1$ the cure probability for stage 1–2 and $\theta_2$ the cure probability for stage 3. The number of treated ($T_{Y_s,i}$), cured ($C_{Y_s,i}$), and remaining infected $R_{Y_s,i}$ in age group $i$ and stage $s$ were:

$$\begin{aligned}
T_{Y_1,i} &= p_{Y_1} Y_{1,i}^-, & C_{Y_1,i} &= \theta_1 T_{Y_1,i}, & R_{Y_1,i} &= (1-\theta_1) T_{Y_1,i},\\
T_{Y_2,i} &= p_{Y_2} Y_{2,i}^-, & C_{Y_2,i} &= \theta_1 T_{Y_2,i}, & R_{Y_2,i} &= (1-\theta_1) T_{Y_2,i},\\
T_{Y_3,i} &= p_{Y_3} Y_{3,i}^-, & C_{Y_3,i} &= \theta_2 T_{Y_3,i}, & R_{Y_3,i} &= (1-\theta_2) T_{Y_3,i}.
\end{aligned}$$

The total number of successfully cured cases for each disease stage was:

$$\begin{aligned}
Y_{1,i}^+ &= Y_{1,i}^- - \theta_1 T_{J1,i},\\
Y_{2,i}^+ &= Y_{2,i}^- - \theta_1 T_{J2,i},\\
Y_{3,i}^+ &= Y_{3,i}^- - \theta_2 T_{J3,i},\\
Y_{u,i}^+ &= Y_{u,i}^- + C_{J1,i} + C_{J2,i} + C_{J3,i}.
\end{aligned}$$

#### Ageing

Let $Y_{s,i}(t)$ denote the population in compartment $Y \in \{D, J, X\}$, stage $s \in \{u, 0, \ldots, 6\}$, and age group $i$ at time $t$. Ageing was modelled by shifting one-fifth of individuals from each age group into the next age group:

$$
\begin{aligned}
Y_{s,i}(t^+) &= Y_{s,i}(t^-) - \frac{1}{5}Y_{s,i}(t^-),\\
Y_{s,i+1}(t^+) &= Y_{s,i+1}(t^-) + \frac{1}{5}Y_{s,i}(t^-), i = 1,\cdots,8\\
Y_{s,9}(t^+) &= Y_{s,9}(t^-) + \frac{1}{5}Y_{s,8}(t^-).
\end{aligned}
$$

where $t^+$ and $t^-$ denote the population immediately before and after the ageing step, respectively.

**Parameter estimation**

We employed Markov chain Monte Carlo (McMC) algorithms to estimate age-specific scaling factors by fitting model-based age-specific HCV prevalence to observed data. These factors ($\boldsymbol{p} = (p_1, \cdots, p_9)$) were assigned uniform prior distributions. Let $\hat{\pi}_i(t; p)$ denote the model-predicted HCV prevalence for age group $i$ at time $t$, and $\pi_i(t)$ the corresponding observation. We constructed the log-likelihood function using the cross-entropy loss:

$$\ell(\boldsymbol{p}) = \sum_{i,t} \pi_i(t) \times \log \hat{\pi}_i(t; \boldsymbol{p}) + (1 - \pi_i(t)) \times \log (1 - \hat{\pi}_i(t; \boldsymbol{p})),$$

The optimal jumping step size was determined through a grid search. The MCMC algorithm was run for 110,000 iterations, with the first 10,000 iterations discarded as burn-in. Convergence was assessed using effective sample size ($> 1{,}000$) and visual inspection of trace plots of posterior samples. To initialize the simulation, the compartmental model was run to equilibrium, and the resulting steady values were used as the initial conditions for all time-varying quantities.

### Main analysis results

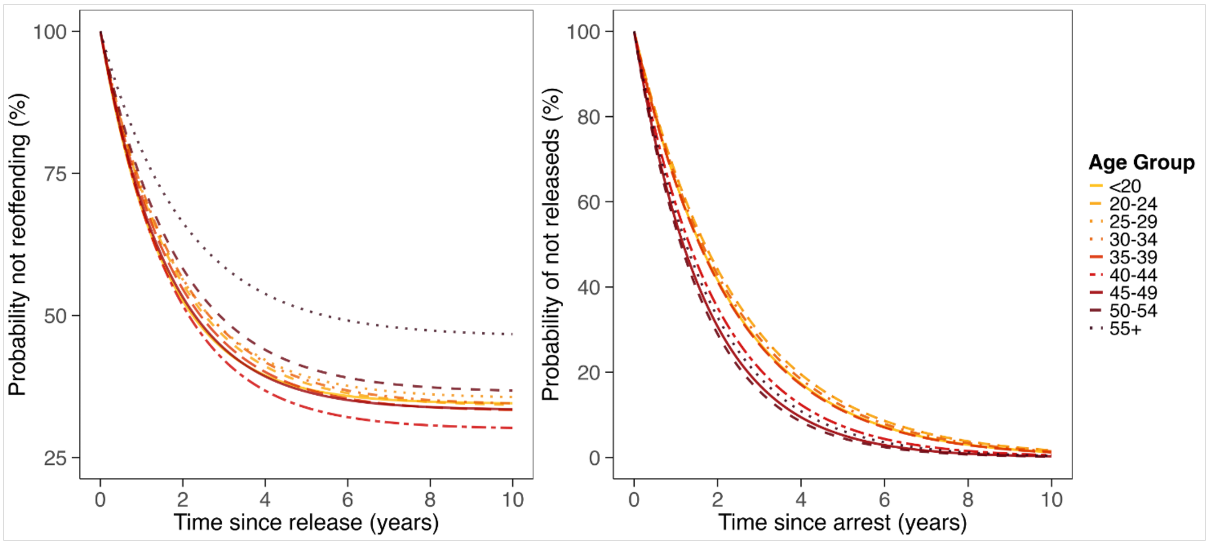

**Figure S1.** Inferred survival curves for recidivism (left) and time spent in prison (right) for sojourns in different age groups.

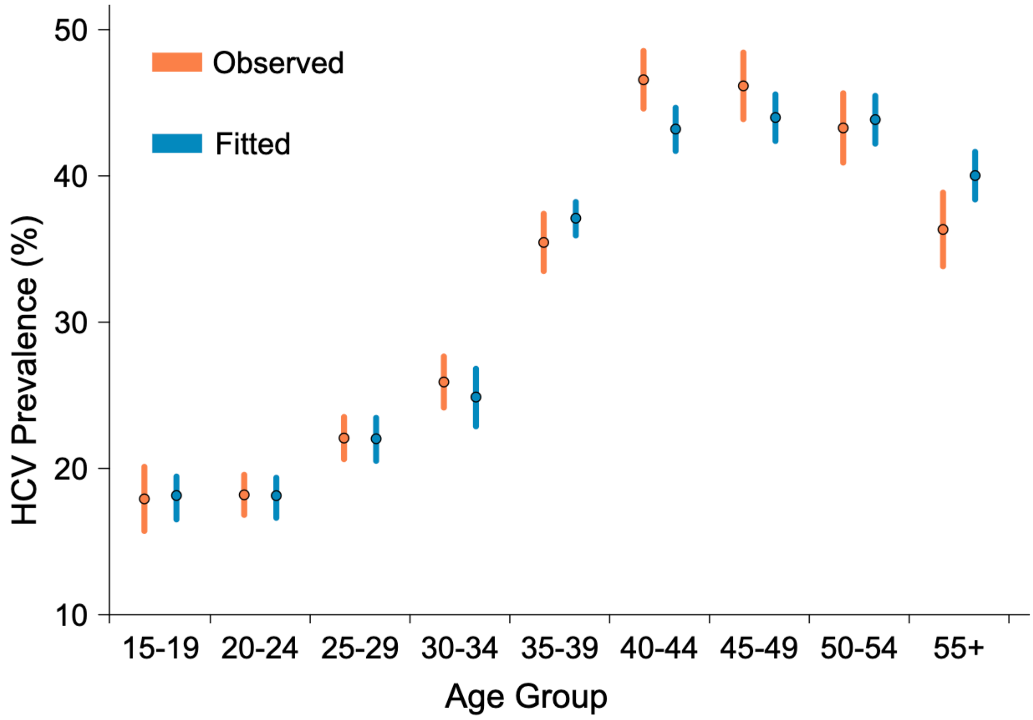

**Figure S2.** Age-specific prevalence of HCV infection among 5,086 PWIDs. Orange dots and lines (left) denote the observed prevalence and corresponding 95% condifence intervals, while blue dots and lines (right) represent the model-estimated posterior medians and 95% credible intervals.

**Table S1.** Percentage reduction (%) in the number of infections by age group under selected treatment strategies over 50 years.

| **Age group (years)** | **PWID (15%)** | **Prisoners (all)** | **Ex-PWID (15%)** | **Combined (12%, all, 13%)** |
|---|---|---|---|---|
| **<20** | 98.89 (98.04–99.16) | 84.78 (63.91–95.88) | 2.56 (1.35–4.65) | 99.90 (99.65–99.98) |
| **20–24** | 98.77 (97.89–99.04) | 88.33 (70.88–96.56) | 10.19 (5.56–14.59) | 99.92 (99.73–99.98) |
| **25–29** | 98.40 (97.36–98.68) | 90.87 (74.59–97.11) | 20.29 (12.74–26.70) | 99.94 (99.78–99.98) |
| **30–34** | 97.83 (96.50–98.22) | 91.02 (75.88–97.03) | 28.70 (18.86–38.19) | 99.95 (99.78–99.98) |
| **35–39** | 97.02 (95.08–97.49) | 90.00 (75.72–96.47) | 33.02 (23.75–45.59) | 99.94 (99.78–99.98) |
| **40–44** | 95.62 (92.98–96.38) | 90.91 (81.67–96.64) | 40.17 (31.27–54.63) | 99.95 (99.84–99.98) |
| **44–49** | 93.38 (89.84–94.88) | 91.17 (84.80–93.95) | 49.15 (39.01–62.53) | 99.96 (99.88–99.98) |
| **50–54** | 90.18 (85.55–92.71) | 90.04 (84.68–91.67) | 57.67 (47.69–68.76) | 99.96 (99.91–99.97) |
| **55+** | 55.15 (48.01–61.55) | 59.18 (55.22–62.60) | 77.86 (69.78–84.73) | 99.88 (99.86–99.89) |

### Sensitivity analysis results

**Table S2.** Estimated prevalence and cumulative number of HCV-related complications, treatments required, and deaths under selected treatment scenarios by year 30.

| | Baseline | Treated group | | | |
|---|---|---|---|---|---|
| | No treatment | PWID (15%) | Prisoners (all) | Ex-PWID (15%) | Combined (12%, all, 13%) |
| **Prevalence (%, by Year 30)** | | | | | |
| **Total** | 21.26 | 10.26 | 9.54 | 9.53 | 0.60 |
| | (18.58–25.08) | (9.59–10.92) | (8.73–11.86) | (7.44–13.15) | (0.51–0.90) |
| **PWID** | 30.31 | 3.10 | 7.61 | 30.31 | 0.70 |
| | (24.92–38.52) | (2.18–4.53) | (5.12–14.75) | (24.92–38.52) | (0.43–1.67) |
| **Prisoners** | 30.19 | 3.87 | 2.77 | 30.19 | 0.29 |
| | (22.17–40.21) | (2.62–5.78) | (1.49–6.02) | (22.17–40.21) | (0.15-0.77) |
| **Ex-PWID** | 18.47 | 12.44 | 10.50 | 3.27 | 0.60 |
| | (16.95–20.05) | (12.00–13.07) | (9.95–11.46) | (2.65–3.96) | (0.56–0.68) |
| **Count (n, by Year 30)** | | | | | |
| **Decompensated cirrhosis** | 7643 | 6567 | 6440 | 4046 | 3081 |
| | (7571–7733) | (6494–6620) | (6363–6576) | (3966–4172) | (3060–3112) |
| **Hepatocellular carcinoma** | 720 | 659 | 652 | 500 | 443 |
| | (716–724) | (655–662) | (647–660) | (496–507) | (441–445) |
| **Treatments** | 0 | 14305 | 18817 | 35802 | 42324 |
| | | (12720–17325) | (16036–22419) | (33784–37110) | (40771-45253) |
| **Deaths** | 8716 | 7675 | 7563 | 4983 | 4004 |
| | (8652–8785) | (7606–7729) | (7482–7701) | (4913–5091) | (3978–4040) |
